# Standardized Comparison of Clinical, Cognitive, Genetic, Neuroimaging, and Fluid Biomarkers for Predicting 24-Month Progression from Mild Cognitive Impairment to Alzheimer’s Disease

**DOI:** 10.64898/2026.08.03.26359630

**Authors:** Sophie Choe

## Abstract

Identifying individuals with mild cognitive impairment (MCI) likely to progress to Alzheimer’s disease (AD) is important for patient management and clinical trial enrollment. Although cognitive assessments, genetics, neuroimaging, and fluid biomarkers are each associated with disease progression, their predictive value has not been systematically compared using an identical cohort and evaluation framework. This study compared the predictive discrimination of clinical, cognitive, genetic, imaging, and cerebrospinal fluid (CSF) biomarkers, individually and combined, for 24-month progression from MCI to AD.

A retrospective analysis used data from 2,430 participants with MCI enrolled in ADNI, including 547 who progressed to AD within 24 months and 1,883 who remained stable. Seven models were evaluated using identical preprocessing and modeling procedures: a clinical baseline (age and sex), the baseline plus a single modality — cognitive assessment, APOE ε4 genotype, structural MRI, CSF biomarkers, or PET biomarkers — and a multimodal model combining all five. Performance was assessed using repeated 5×10 stratified cross-validation. Out-of-fold predictions from a separate 5-fold split were used to estimate confidence intervals and compare AUCs via DeLong’s test with Holm–Bonferroni correction.

Discrimination increased progressively across modalities. The clinical baseline achieved an AUC of 0.556; adding APOE ε4 genotype increased performance to 0.692, CSF biomarkers to 0.729, PET biomarkers to 0.783, structural MRI to 0.836, and cognitive assessment to 0.918. Cognitive assessment significantly outperformed all other individual modalities, including MRI (ΔAUC = 0.079, P < 0.001). The multimodal model achieved the highest overall discrimination (AUC = 0.933), significantly outperforming cognitive assessment alone (ΔAUC = 0.016, P < 0.001), though it required complete data from only 20.5% of participants, versus 99.3% for cognitive assessment.

Within a common evaluation framework, cognitive assessment demonstrated the greatest predictive discrimination among individual modalities for 24-month progression from MCI to AD, followed by structural MRI and PET. A multimodal model achieved the highest overall discrimination but required complete data from only one-fifth of the cohort. These findings suggest that routinely collected cognitive assessments capture substantial prognostic information, while full multimodal integration offers only modest incremental value relative to its reduced applicability.

## Background

Alzheimer’s disease (AD) is the leading cause of dementia worldwide and is characterized by a prolonged preclinical phase followed by progressive cognitive and functional decline [1,2]. Individuals with mild cognitive impairment (MCI) represent a heterogeneous population: while some progress to AD within a few years, others remain clinically stable or develop alternative neurodegenerative disorders [3,4]. Accurately identifying individuals at greatest risk of progression is therefore important for patient counseling, prognosis, clinical trial enrollment, and the efficient use of increasingly available disease-modifying therapies and biomarker testing.

A wide range of biomarkers has been investigated for predicting progression from MCI to AD [5-8]. Cognitive assessments remain the cornerstone of clinical evaluation because they directly measure the functional consequences of neurodegeneration and are inexpensive, non-invasive, and widely available [9–11]. Genetic risk factors, particularly the apolipoprotein E (APOE) ε4 allele, are associated with increased susceptibility to AD but provide limited information regarding the timing of clinical progression [12,13]. Structural magnetic resonance imaging (MRI) quantifies regional brain atrophy [14], positron emission tomography (PET) visualizes amyloid and glucose metabolism [15], and cerebrospinal fluid (CSF) biomarkers reflect the underlying molecular pathology of AD [16]. Together, these modalities provide complementary information regarding disease risk and progression.

Numerous studies have reported predictive models based on individual biomarker modalities or multimodal combinations [5–8]. However, direct comparison of their reported performance remains difficult because studies frequently differ in participant selection, outcome definitions, follow-up duration, preprocessing strategies, feature engineering, machine learning algorithms, validation procedures, and performance metrics. Consequently, higher reported performance for one biomarker modality may reflect methodological differences rather than superior prognostic value. This lack of standardized evaluation has made it difficult to determine which routinely available biomarkers provide the greatest predictive discrimination for progression from MCI to AD.

To address this gap, we performed a standardized comparison of commonly used clinical, cognitive, genetic, neuroimaging, and fluid biomarker modalities using a single cohort from the Alzheimer’s Disease Neuroimaging Initiative (ADNI) [17]. All models were developed using an identical machine learning pipeline, including the same preprocessing strategy, feature engineering, model architecture, outcome definition, and repeated cross-validation framework. By controlling these methodological factors, differences in predictive performance more accurately reflect the relative prognostic value of each biomarker modality rather than differences in analytical approach.

We hypothesized that cognitive assessment would provide greater predictive discrimination than genetic, neuroimaging, or fluid biomarkers for predicting 24-month progression from MCI to AD. Establishing the relative predictive value of these routinely available modalities may help guide clinical assessment, optimize participant selection for clinical trials, and inform the development of practical risk prediction tools for Alzheimer’s disease.

## Methods

### Study design and objective

This retrospective cohort study compared the predictive performance of clinical characteristics, cognitive assessment, genetic information, structural magnetic resonance imaging (MRI), cerebrospinal fluid (CSF) biomarkers, and positron emission tomography (PET) biomarkers for predicting progression from mild cognitive impairment (MCI) to Alzheimer’s disease within 24 months. The primary objective was to quantify and compare the discriminatory performance of each biomarker modality using an identical participant cohort, preprocessing pipeline, machine learning algorithm, and evaluation framework, thereby isolating the contribution of the data modality from differences in model architecture or optimization.

### Data source and participants

Data were obtained from the Alzheimer’s Disease Neuroimaging Initiative (ADNI), a longitudinal multicenter observational study established to identify biomarkers for the early detection and monitoring of Alzheimer’s disease [17].

Participants with a baseline diagnosis of MCI and a known clinical diagnosis at 24 months were eligible for inclusion. The prediction target was progression to clinically diagnosed Alzheimer’s disease within 24 months of baseline assessment. Participants who converted to Alzheimer’s disease were classified as converters, whereas those who remained free of an Alzheimer’s disease diagnosis were classified as non-converters.

The final study cohort comprised 2,430 participants, including 547 converters (22.5%) and 1,883 non-converters (77.5%).

### Predictor variables

Seven prediction models were evaluated. The baseline model included age and sex. Five additional models evaluated the predictive performance of individual biomarker modalities. A seventh, multimodal model combined clinical variables with all five biomarker modalities to evaluate the incremental predictive value of integrating multiple data sources.

The evaluated modalities were:

**Cognitive assessment:** Mini-Mental State Examination (MMSE) [18], Logical Memory delayed recall (LDELTOTAL) [19], and Rey Auditory Verbal Learning Test immediate recall (RAVLT immediate) [20]. Logical Memory delayed recall was selected as the representative episodic memory measure based on superior discriminative performance relative to immediate recall (LIMMTOTAL) in preliminary comparison (AUC 0.918 vs. 0.908).

**Genetic information:** apolipoprotein E (APOE) ε4 allele count [12,13].

**Structural MRI:** hippocampal volume, entorhinal cortex volume, middle temporal gyrus volume, whole brain volume, and ventricular volume [14,21].

**CSF biomarkers:** amyloid-β42 (Aβ42), total tau, and phosphorylated tau [22].

**PET biomarkers:** florbetapir (AV45) standardized uptake value ratio (SUVR) and fluorodeoxyglucose (FDG) SUVR [23].

No additional feature engineering or feature selection was performed.

### Data preprocessing

All preprocessing was performed independently within each training fold to prevent information leakage. Preprocessing and model fitting were implemented as a single scikit-learn pipeline so that all preprocessing parameters were estimated exclusively from the training data within each fold.

Continuous variables were imputed using the median value calculated from the corresponding training fold. Categorical variables were imputed using the most frequent category and one-hot encoded. The fitted preprocessing pipeline was then applied unchanged to the corresponding validation fold. Participants were not excluded because of missing predictor values in the primary analyses.

Missingness varied substantially across modalities, ranging from less than 1% for demographic and cognitive variables to more than 50% for CSF and PET biomarkers (Table 2). Because of this variation, complete-case sensitivity analyses were performed for the CSF, PET, and multimodal models.

### Machine learning model

All experiments used the same Extreme Gradient Boosting (XGBoost) binary classifier [24] to ensure that observed differences in predictive performance reflected differences in biomarker modality rather than differences in model architecture.

Class imbalance was addressed by calculating the ratio of non-converters to converters within each training fold and applying this value as the scale_pos_weight parameter during model training. Model hyperparameters were fixed across all experiments (300 trees, learning rate 0.03, maximum depth 4, minimum child weight 3, subsample 0.8, column subsampling 0.8, gamma 0.1, L2 regularization = 1.0). No experiment-specific hyperparameter optimization was performed.

### Model evaluation

Model performance was estimated using repeated stratified five-fold cross-validation with ten repetitions, yielding 50 independent validation folds [25].

Within each training fold, the optimal probability threshold was determined by maximizing the Youden index [26]. The selected threshold was then applied unchanged to the corresponding validation fold.

The primary performance metric was the area under the receiver operating characteristic curve (AUC) [27]. Secondary performance measures included accuracy, balanced accuracy [28], sensitivity, specificity, precision, negative predictive value, F1 score, and Brier score [29].

Mean performance and standard deviations were calculated across all validation folds.

### Out-of-fold prediction analysis

To enable paired statistical comparison between models, an additional stratified five-fold cross-validation was performed to generate one out-of-fold prediction for every participant. These predictions were used for receiver operating characteristic analysis and pairwise statistical testing. Because this out-of-fold analysis used a single five-fold split rather than the repeated cross-validation used for primary performance estimation, AUC values derived from this analysis may differ slightly from the repeated cross-validation estimates reported as primary results. Ninety-five percent confidence intervals for AUC were estimated using 2,000 bootstrap resamples [30].

### Sensitivity analysis

Because CSF, PET, and multimodal predictor sets exhibited substantial missingness, complete-case sensitivity analyses were performed for these three models [31]. For each model, the primary imputed model was first evaluated on the subset of participants with complete, non-imputed data for all predictors in that feature set. A second model was then trained and evaluated exclusively on that complete-case subset using the identical cross-validation procedure. Because both models were evaluated on the same participants with identical outcome labels and fold assignments, predictive performance was compared using paired DeLong tests [32]. This analysis assessed whether median imputation materially affected predictive performance among participants with observed data.

### Statistical analysis

Baseline characteristics were compared between converters and non-converters using Welch’s t-test [33] for continuous variables and Pearson’s chi-square test [34] for categorical variables. Standardized mean differences (Cohen’s d) [35] were calculated as complementary measures of effect size for continuous variables.

Differences in AUC between prediction models were evaluated using paired DeLong tests for correlated receiver operating characteristic curves based on paired out-of-fold predictions. Because multiple pairwise comparisons were performed, family-wise error was controlled using the Holm–Bonferroni procedure [36]. Two-sided adjusted P values less than 0.05 were considered statistically significant.

No a priori power calculation was performed. Sample size was determined by the number of eligible participants available in ADNI with a baseline MCI diagnosis and known clinical diagnosis at 24 months, consistent with the retrospective, secondary-data nature of this study.

All analyses were performed in Python using scikit-learn and XGBoost.

## Results

### Study Cohort

The final study cohort comprised 2,430 participants with a baseline diagnosis of MCI and a known clinical diagnosis at 24 months, including 547 converters (22.5%) and 1,883 non-converters (77.5%). Baseline characteristics differed significantly between converters and non-converters across nearly all measured domains (Table 1). Converters were older than non-converters (74.34 vs. 72.48 years, P<0.001, SMD=0.25) and were more likely to be male (58.0% vs. 50.8%, P=0.004). Cognitive assessment showed the largest standardized differences of any domain: MMSE (SMD=1.77), Logical Memory delayed recall (SMD=1.64), and RAVLT immediate recall (SMD=1.47) were all substantially lower among converters. APOE ε4 allele count was higher among converters (0.86 vs. 0.46, SMD=0.63). Structural MRI measures showed consistently smaller volumes among converters, most notably hippocampal volume (SMD=1.28) and entorhinal cortex volume (SMD=1.09). CSF biomarkers showed the expected Alzheimer’s disease pathological pattern among converters — lower amyloid-β42 and higher total and phosphorylated tau (SMD=0.80–0.85) — and PET biomarkers showed higher amyloid burden (florbetapir SUVR, SMD=1.13) and lower glucose metabolism (FDG SUVR, SMD=1.32).

**Table 1.** Cohort characteristics by 24-month progression status.

| Characteristic | Non-converters | Converters | P value | SMD |
| --- | --- | --- | --- | --- |
| **Demographics** |  |  |  |  |
| Age, years, mean (SD) [N] | 72.48 (7.24) [1879] | 74.34 (7.67) [547] | <0.001 | 0.25 |
| Sex, female, n (%) | 927 (49.2%) | 230 (42.0%) | 0.004 | — |
| **Cognitive assessment** |  |  |  |  |
| MMSE, mean (SD) [N] | 28.23 (1.96) [1882] | 24.46 (2.65) [547] | <0.001 | 1.77 |
| Logical Memory delayed recall, mean (SD) [N] | 9.44 (4.87) [1879] | 2.13 (2.65) [547] | <0.001 | 1.64 |
| RAVLT immediate recall, mean (SD) [N] | 40.28 (11.67) [1875] | 24.24 (7.54) [544] | <0.001 | 1.47 |
| **Genetics** |  |  |  |  |
| APOE ε4 allele count, mean (SD) [N] | 0.46 (0.62) [1680] | 0.86 (0.69) [533] | <0.001 | 0.63 |
| **Structural MRI** |  |  |  |  |
| Hippocampal volume, mm<sup>3</sup>, mean (SD) [N] | 7221.99 (1031.27) [1624] | 5892.04 (1063.65) [457] | <0.001 | 1.28 |
| Entorhinal cortex volume, mm<sup>3</sup>, mean (SD) [N] | 3810.53 (742.64) [1609] | 2991.52 (777.68) [443] | <0.001 | 1.09 |
| Middle temporal gyrus volume, mm<sup>3</sup>, mean (SD) [N] | 20427.31 (2871.74) [1609] | 17572.63 (3012.46) [443] | <0.001 | 0.98 |
| Whole brain volume, cm<sup>3</sup>, mean (SD) [N] | 1040.38 (107.14) [1763] | 984.90 (117.46) [529] | <0.001 | 0.51 |
| Ventricular volume, cm<sup>3</sup>, mean (SD) [N] | 36.32 (20.91) [1736] | 49.11 (23.88) [516] | <0.001 | 0.59 |
| **CSF biomarkers** |  |  |  |  |
| Amyloid-β42, pg/mL, mean (SD) [N] | 936.38 (371.15) [703] | 648.24 (256.59) [320] | <0.001 | 0.85 |
| Total tau, pg/mL, mean (SD) [N] | 260.28 (114.81) [878] | 357.22 (138.93) [332] | <0.001 | 0.80 |
| Phosphorylated tau, pg/mL, mean (SD) [N] | 24.68 (12.83) [877] | 35.49 (15.12) [332] | <0.001 | 0.80 |
| **PET biomarkers** |  |  |  |  |
| Florbetapir (AV45) SUVR, mean (SD) [N] | 1.16 (0.21) [922] | 1.40 (0.22) [211] | <0.001 | 1.13 |
| FDG-PET SUVR, mean (SD) [N] | 1.24 (0.14) [1141] | 1.06 (0.14) [374] | <0.001 | 1.32 |

SMD, standardized mean difference (absolute value of Cohen's *d*); not computed for categorical variables. Male sex is the complement of female sex and is omitted to avoid redundancy. *P* values from Welch's *t*-test (continuous variables) or Pearson's chi-square test (categorical variables). Sample sizes vary by variable due to missing data.

Missingness varied substantially by modality (Table 2). Demographic and cognitive variables were nearly complete (>99% availability), APOE genotype was available for 91.1% of participants, and structural MRI measures were available for 79.9–84.6% of participants. CSF and PET biomarkers showed the greatest missingness, with complete data available for 37.7–57.9% of participants depending on the specific biomarker. Consequently, only 499 participants (20.5%) had complete data across all modalities required for the multimodal model.

**Table 2.** Missingness by variable across all modalities.

| Modality | Variable | N available | N missing | % missing |
| --- | --- | --- | --- | --- |
| Demographics | Age, years | 2426 | 4 | 0.2% |
|  | Sex | 2430 | 0 | 0.0% |
| Cognitive assessment | MMSE | 2429 | 1 | 0.0% |
|  | Logical Memory delayed recall (LDELTOTAL) | 2426 | 4 | 0.2% |
|  | RAVLT immediate recall | 2419 | 11 | 0.5% |
| Genetics | APOE $\epsilon 4$ allele count | 2213 | 217 | 8.9% |
| Structural MRI | Hippocampal volume, mm<sup>3</sup> | 2081 | 349 | 14.4% |
|  | Entorhinal cortex volume, mm<sup>3</sup> | 2052 | 378 | 15.6% |
|  | Middle temporal gyrus volume, mm<sup>3</sup> | 2052 | 378 | 15.6% |
|  | Whole brain volume, mm<sup>3</sup> | 2292 | 138 | 5.7% |
|  | Ventricular volume, mm<sup>3</sup> | 2252 | 178 | 7.3% |
| CSF biomarkers | Amyloid-β42, pg/mL | 1023 | 1407 | 57.9% |
|  | Total tau, pg/mL | 1210 | 1220 | 50.2% |
|  | Phosphorylated tau, pg/mL | 1209 | 1221 | 50.2% |
| PET biomarkers | Florbetapir (AV45) SUVR | 1133 | 1297 | 53.4% |
|  | FDG-PET SUVR | 1515 | 915 | 37.7% |

*N total = 2,430. Percentages rounded to one decimal place.*

### Comparison of Individual Modalities

The predictive performance of each modality is summarized in Table 3. The baseline clinical model, comprising age and sex alone, achieved an AUC of 0.556 (accuracy=0.539), indicating minimal discrimination between converters and non-converters in the absence of any biomarker information. Among individual modalities evaluated in combination with clinical variables, cognitive assessment achieved the highest discriminatory performance (AUC=0.918, accuracy=0.847, sensitivity=0.857, specificity=0.845, Brier score=0.110). Structural MRI achieved the second-highest performance (AUC=0.836), followed by PET biomarkers (AUC=0.783), CSF biomarkers (AUC=0.729), and APOE genotype (AUC=0.692). The multimodal model, integrating all five modalities with clinical variables, achieved the highest overall performance (AUC=0.933, accuracy=0.876, sensitivity=0.818, specificity=0.893, Brier score=0.094).

**Table 3.** Predictive performance by modality (primary 5×10 repeated cross-validation)

| Model | N | AUC | Accuracy | Sensitivity | Specificity | Brier score |
| --- | --- | --- | --- | --- | --- | --- |
| Clinical only | 2430 | 0.556 | 0.539 | 0.540 | 0.538 | 0.244 |
| Clinical + APOE | 2430 | 0.692 | 0.634 | 0.664 | 0.626 | 0.216 |
| Clinical + CSF | 2430 | 0.729 | 0.694 | 0.578 | 0.728 | 0.195 |
| Clinical + PET | 2430 | 0.783 | 0.699 | 0.676 | 0.706 | 0.181 |
| Clinical + MRI | 2430 | 0.836 | 0.773 | 0.732 | 0.785 | 0.151 |
| Clinical + Cognitive | 2430 | 0.918 | 0.847 | 0.857 | 0.845 | 0.110 |
| **Multimodal** | 2430 | **0.933** | 0.876 | 0.818 | 0.893 | 0.094 |

*Values represent means across 50 validation folds (5-fold cross-validation × 10 repetitions). N reflects total participants entering each experiment (missing predictor values were imputed; see Methods).*

Cognitive assessment improved AUC by **0.362** compared with the clinical baseline, whereas MRI improved AUC by **0.280**, PET by **0.227**, CSF by **0.173**, and APOE by **0.136**.

Notably, although the multimodal model achieved the highest AUC and specificity, its sensitivity (0.818) was lower than that of cognitive assessment alone (0.857), reflecting a shift in the optimal classification threshold under the multimodal model’s score distribution (Figure 1).

**Figure 1.**
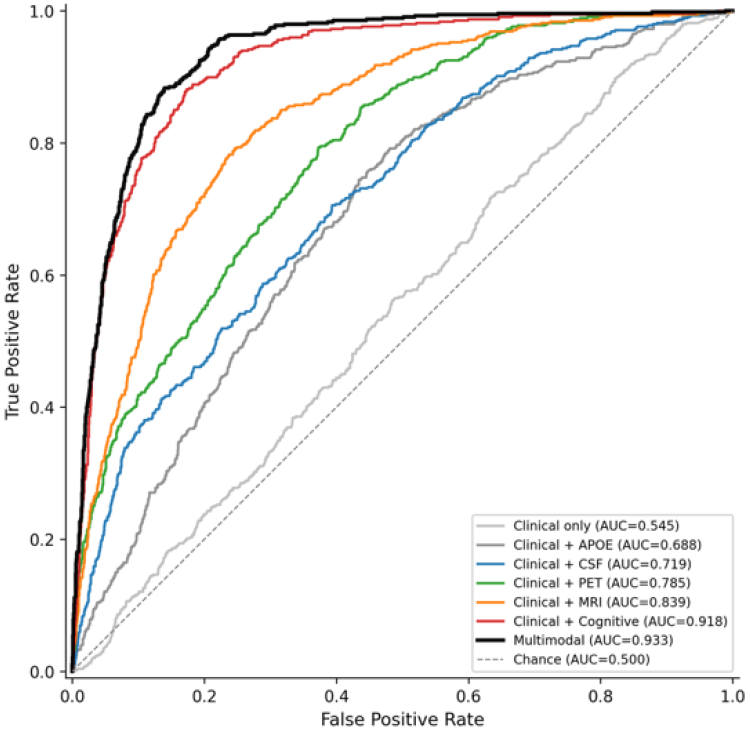
Receiver operating characteristic curves for individual and multimodal prediction models. Receiver operating characteristic (ROC) curves for out-of-fold predictions from all seven prediction models. The diagonal dashed line represents chance-level discrimination (AUC = 0.5). AUC values for each model are shown in the legend. See Table 3 for primary performance metrics and Table 4 for pairwise statistical comparisons between models.

### Pairwise Statistical Comparisons

Pairwise DeLong tests were performed on paired out-of-fold predictions to compare discriminatory performance between all seven models (21 comparisons total; Table 4). All comparisons remained statistically significant after Holm–Bonferroni correction for multiple testing (all adjusted P<0.05). The clinical-only model was significantly outperformed by every other model, including APOE genotype, the weakest individual biomarker modality (ΔAUC=−0.143, P<0.001). Cognitive assessment significantly outperformed every other single modality, including structural MRI (ΔAUC=0.079, P<0.001), PET biomarkers (ΔAUC=0.133, P<0.001), CSF biomarkers (ΔAUC=0.198, P<0.001), and APOE genotype (ΔAUC=0.230, P<0.001).

**Table 4.**
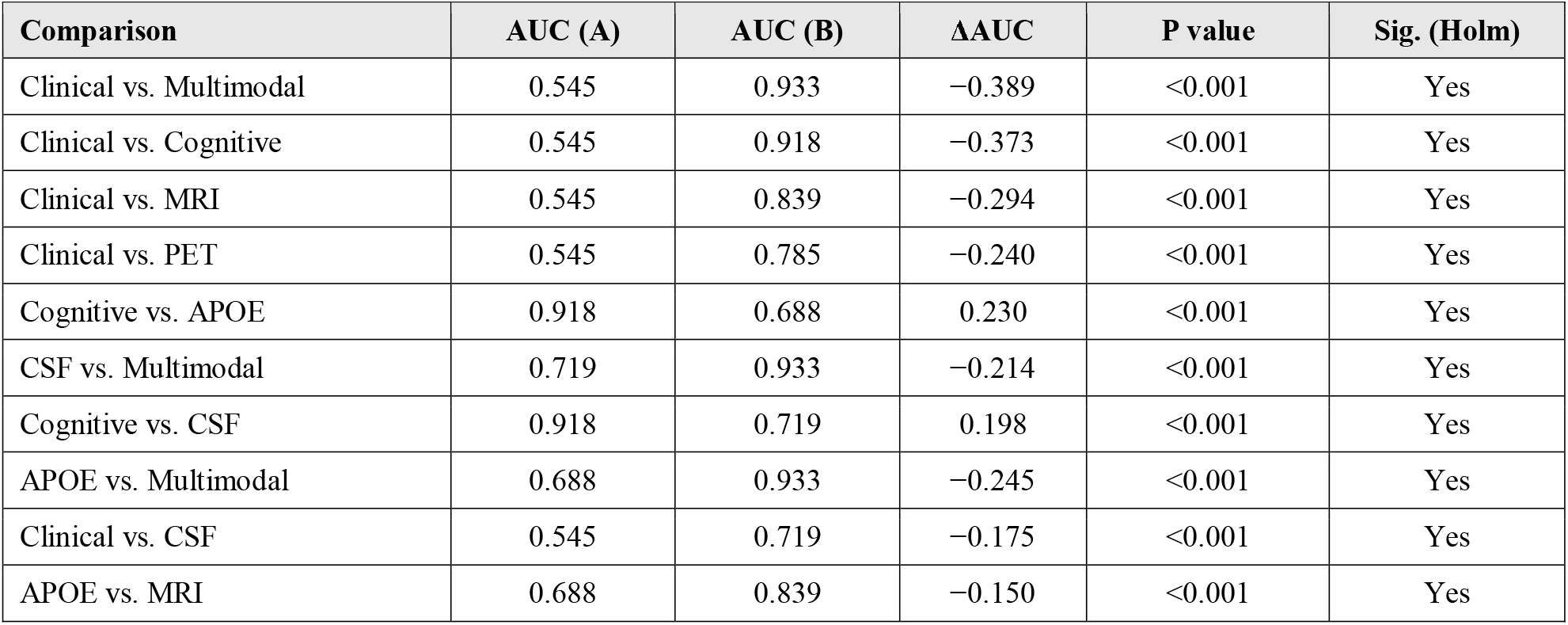
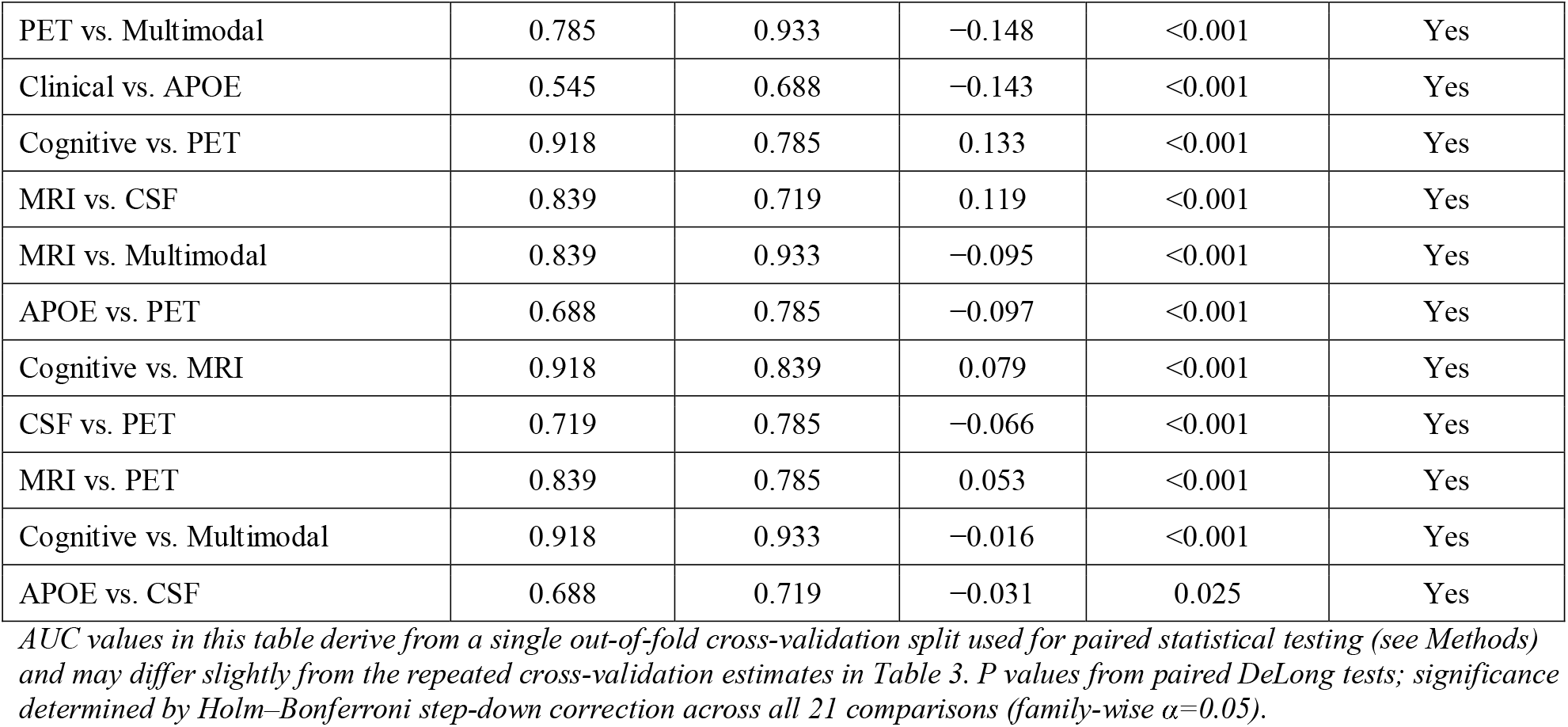
Pairwise DeLong comparisons between all models (out-of-fold predictions)

| Comparison | AUC (A) | AUC (B) | $\Delta AUC$ | P value | Sig. (Holm) |
| --- | --- | --- | --- | --- | --- |
| Clinical vs. Multimodal | 0.545 | 0.933 | -0.389 | <0.001 | Yes |
| Clinical vs. Cognitive | 0.545 | 0.918 | -0.373 | <0.001 | Yes |
| Clinical vs. MRI | 0.545 | 0.839 | -0.294 | <0.001 | Yes |
| Clinical vs. PET | 0.545 | 0.785 | -0.240 | <0.001 | Yes |
| Cognitive vs. APOE | 0.918 | 0.688 | 0.230 | <0.001 | Yes |
| CSF vs. Multimodal | 0.719 | 0.933 | -0.214 | <0.001 | Yes |
| Cognitive vs. CSF | 0.918 | 0.719 | 0.198 | <0.001 | Yes |
| APOE vs. Multimodal | 0.688 | 0.933 | -0.245 | <0.001 | Yes |
| Clinical vs. CSF | 0.545 | 0.719 | -0.175 | <0.001 | Yes |
| APOE vs. MRI | 0.688 | 0.839 | -0.150 | <0.001 | Yes |
| PET vs. Multimodal | 0.785 | 0.933 | -0.148 | <0.001 | Yes |
| Clinical vs. APOE | 0.545 | 0.688 | -0.143 | <0.001 | Yes |
| Cognitive vs. PET | 0.918 | 0.785 | 0.133 | <0.001 | Yes |
| MRI vs. CSF | 0.839 | 0.719 | 0.119 | <0.001 | Yes |
| MRI vs. Multimodal | 0.839 | 0.933 | -0.095 | <0.001 | Yes |
| APOE vs. PET | 0.688 | 0.785 | -0.097 | <0.001 | Yes |
| Cognitive vs. MRI | 0.918 | 0.839 | 0.079 | <0.001 | Yes |
| CSF vs. PET | 0.719 | 0.785 | -0.066 | <0.001 | Yes |
| MRI vs. PET | 0.839 | 0.785 | 0.053 | <0.001 | Yes |
| Cognitive vs. Multimodal | 0.918 | 0.933 | -0.016 | <0.001 | Yes |
| APOE vs. CSF | 0.688 | 0.719 | -0.031 | 0.025 | Yes |

The multimodal model significantly outperformed every individual modality, including cognitive assessment, the strongest single predictor (AUC 0.933 vs. 0.918; ΔAUC=0.016, P<0.001) (Figure 2). However, this incremental improvement was modest in absolute magnitude and was achieved using only 499 of 2,430 participants (20.5%) with complete data across all modalities, compared with 2,413 participants (99.3%) with complete cognitive assessment data.

**Figure 2.**
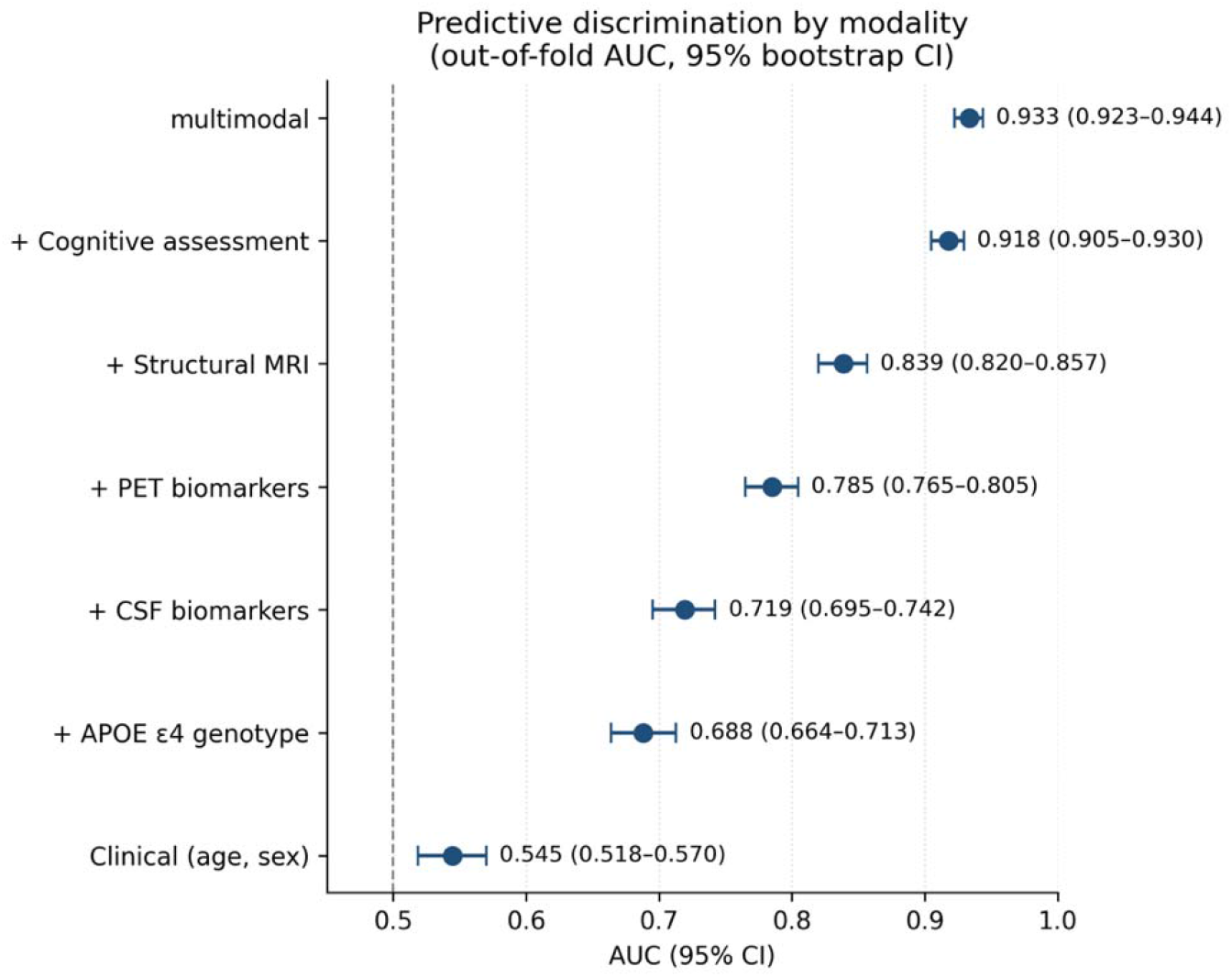
Pairwise differences in discrimination (AUC) across all modality comparisons. Forest plot of pairwise AUC differences (ΔAUC) between all seven prediction models, derived from paired DeLong tests on out-of-fold predictions. All 21 comparisons remained statistically significant after Holm–Bonferroni correction for multiple testing (P<0.05). See Table 4 for full numerical results.

### Sensitivity Analysis for Missing Data

Because CSF, PET, and multimodal predictor sets exhibited substantial missingness, complete-case sensitivity analyses were performed to assess whether median imputation materially affected predictive performance (Table 5). For each of the three models, predictions from the primary imputed model were compared with predictions from a model retrained exclusively on complete cases, using paired DeLong tests.

**Table 5.** Complete-case sensitivity analysis for models with substantial missingness.

| Model | AUC (full, imputed) | N (complete-case) | % complete-case | AUC (imputed model, complete-case subset) | AUC (complete-case, refit) | P value |
| --- | --- | --- | --- | --- | --- | --- |
| Clinical + CSF | 0.719<br>(95% CI 0.695–0.742) | 1017 | 41.9% | 0.758 | 0.770<br>(95% CI 0.741–0.800) | 0.058 |
| Clinical + PET | 0.785<br>(95% CI 0.765–0.805) | 968 | 39.8% | 0.862 | 0.863<br>(95% CI 0.836–0.888) | 0.838 |
| Multimodal | 0.933<br>(95% CI 0.923–0.944) | 499 | 20.5% | 0.940 | 0.943<br>(95% CI 0.924–0.963) | 0.583 |

*N = 2,430 for the full imputed cohort in all three models. AUC (imputed model, complete-case subset): the primary model, trained on the full imputed cohort, evaluated only on participants with complete non-imputed data. AUC (complete-case, refit): a separate model trained and evaluated exclusively on the complete-case subset. P values from paired DeLong tests comparing these two sets of predictions within the same participants.*

For CSF biomarkers, the imputed model applied to the complete-case subset (n=1,017) achieved an AUC of 0.758, compared with an AUC of 0.770 for the model retrained on complete cases only; this difference did not reach statistical significance (P=0.058). For PET biomarkers, the corresponding AUCs were 0.862 and 0.863 (n=968, P=0.838), and for the multimodal model, 0.940 and 0.943 (n=499, P=0.583). In all three cases, complete-case AUCs were modestly higher than the corresponding imputed-model AUCs on the full cohort (Table 3), which may reflect systematic differences between participants with complete biomarker data and those with missing data, rather than distortion introduced by imputation itself. None of the three imputed-versus-complete-case comparisons reached statistical significance, supporting the validity of the primary, imputation-based results reported above.

## Discussion

In this standardized comparison of biomarker modalities for predicting 24-month progression from mild cognitive impairment (MCI) to Alzheimer’s disease (AD), three principal findings emerged. First, cognitive assessment demonstrated the strongest predictive discrimination among all individual modalities. Second, structural magnetic resonance imaging (MRI) was the highest-performing biological modality, outperforming positron emission tomography (PET), cerebrospinal fluid (CSF) biomarkers, and apolipoprotein E (APOE) ε4 genotype. Third, integrating cognitive, genetic, imaging, and fluid biomarkers produced the highest overall predictive performance. We notice, however, that the incremental improvement over cognitive assessment alone was modest (ΔAUC=0.016) and accompanied by substantially reduced data availability (missingness 0.5% vs 57.9%). Together, these findings provide a quantitative benchmark for comparing commonly used prognostic modalities under an identical analytical framework.

A major challenge in the Alzheimer’s disease prediction literature is that reported model performance is rarely directly comparable. Previous studies have evaluated different participant populations, follow-up periods, outcome definitions, preprocessing strategies, machine learning algorithms, and validation procedures, making it difficult to determine whether differences in reported performance reflect the intrinsic value of a biomarker or methodological variation [5–8]. Few studies have directly compared these modalities within the same participant cohort using a common analytical framework. By evaluating all modalities within the same ADNI cohort using identical preprocessing, feature engineering, model architecture, outcome definition, and repeated cross-validation, the present study minimizes methodological confounding and enables a fair comparison of the relative prognostic information contained within each modality.

The superior performance of cognitive assessment, seen in this study, is both biologically and clinically plausible. Cognitive tests measure the cumulative functional consequences of multiple pathological processes, integrating the downstream effects of amyloid deposition, tau pathology, neurodegeneration, cerebrovascular disease, cognitive reserve, and other factors that ultimately determine clinical impairment [9–11]. In contrast, individual biological biomarkers typically quantify a single aspect of disease biology, such as genetic susceptibility, amyloid accumulation, tau pathology, or structural neurodegeneration [12– 15]. Consequently, although biological biomarkers provide essential mechanistic information and play a central role in the biological diagnosis, staging, and increasingly the therapeutic management of Alzheimer’s disease, baseline cognitive performance may more directly reflect the cumulative functional consequences of these pathological processes that determine near-term clinical progression. The superior predictive performance of cognitive assessment observed in this study should therefore be interpreted specifically in the context of prognostic prediction, rather than as evidence that cognitive testing is intrinsically more informative than biological biomarkers for diagnosis. Imaging and fluid biomarkers remain indispensable for establishing Alzheimer’s disease pathology and guiding treatment decisions, whereas cognitive assessment appears to capture downstream manifestations of disease that are particularly valuable for predicting short-term clinical progression.

Among the biological modalities, structural MRI consistently achieved the highest predictive discrimination. MRI-derived measures of regional brain atrophy represent established neurodegeneration and are widely available in routine clinical practice, making MRI an attractive adjunct when cognitive assessment alone is insufficient [14]. PET imaging and CSF biomarkers remain indispensable for biological diagnosis, disease staging, clinical trial enrollment, and therapeutic decision-making [37]. However, biological specificity does not necessarily translate into superior prediction of short-term clinical outcomes. The present findings highlight the distinction between diagnostic biomarkers, which identify Alzheimer’s disease pathology, and prognostic biomarkers, which estimate the likelihood of future clinical decline.

Although multimodal integration achieved the highest overall discrimination, the improvement over cognitive assessment alone was relatively modest (AUC 0.933 versus 0.918, *P* < 0.001). More importantly, this improvement was obtained using only 499 participants (20.5%) with complete multimodal data, whereas cognitive assessment alone was available for 2,413 participants (99.3%). This trade-off between predictive performance, data availability, cost, and assessment burden has important implications for clinical implementation. While comprehensive multimodal models may be appropriate for specialized research settings or clinical trials, cognitive assessment alone provides substantial prognostic information that can be obtained rapidly, inexpensively, and non-invasively in routine clinical practice.

Interpretation of the PET and CSF results should consider the substantial proportion of missing biomarker data (Table 2). Whereas demographic, cognitive, and MRI variables were available for most participants, PET and CSF measurements were unavailable for approximately 40–58% of the cohort. Complete-case sensitivity analyses demonstrated that median imputation did not materially alter predictions for participants with observed biomarker values (Table 5), suggesting that the imputation procedure itself did not introduce measurable bias. Nevertheless, predictive performance improved when analyses were restricted to participants with complete PET, CSF, or multimodal data, indicating that these subsets were not fully representative of the overall cohort. In ADNI, PET imaging was introduced during later study phases and lumbar puncture required separate participant consent, both of which likely contributed to systematic differences between participants with and without biomarker measurements [23]. Accordingly, the observed performance of PET, CSF, and multimodal models should be interpreted within the context of cohort selection and data availability.

The present findings have several practical implications. Cognitive assessment is inexpensive, non-invasive, rapidly administered, and routinely performed in memory clinics worldwide. Its strong predictive performance suggests that it may serve as an effective first-line tool for prognostic risk stratification, with MRI providing additional prognostic information when clinically indicated and PET or CSF biomarkers contributing complementary biological characterization. Rather than replacing biological biomarkers, cognitive assessment and biological measures should be viewed as complementary tools that address different clinical questions: prediction of future cognitive decline versus confirmation of underlying Alzheimer’s disease pathology.

This study has several strengths. All modalities were evaluated within the same participant cohort using a standardized analytical framework, allowing meaningful comparison of predictive performance while minimizing methodological confounding. Repeated stratified cross-validation, out-of-fold evaluation, bootstrap confidence intervals, paired DeLong testing, and sensitivity analyses provided a rigorous statistical assessment of model discrimination and demonstrated that the principal findings were robust to the handling of missing biomarker data. Rather than identifying a single “best” biomarker, these findings suggest a pragmatic approach to risk stratification. Routine cognitive assessment may serve as an efficient first-line prognostic tool, with structural MRI, PET, and CSF biomarkers providing complementary biological information when additional diagnostic certainty or treatment planning is required. Future work should evaluate whether similar modality rankings are observed in more diverse clinical populations and across international cohorts.

Several limitations should be acknowledged. First, ADNI is a highly characterized research cohort and may not fully represent routine clinical populations, making external validation essential before clinical implementation [38]. Second, cognitive assessments contribute to the clinical diagnosis of Alzheimer’s disease and therefore may introduce partial circularity between predictors and outcome. Third, this study evaluated baseline predictors of 24-month progression and did not investigate longitudinal changes in biomarkers or cognition. Finally, although discrimination was comprehensively assessed, additional evaluation of calibration, clinical utility, and decision-curve analysis would further inform implementation in clinical practice.

In conclusion, this standardized comparison demonstrates that cognitive assessment provides the strongest single-modality prediction of 24-month progression from MCI to Alzheimer’s disease, structural MRI is the highest-performing biological modality, and multimodal integration yields the greatest overall discrimination at the expense of substantially reduced data completeness. By evaluating all major biomarker domains using an identical analytical framework, this study establishes a quantitative benchmark for future prognostic studies and provides evidence that routinely collected cognitive assessments capture a remarkable proportion of the prognostic information required for short-term prediction of Alzheimer’s disease progression.

## Conclusions

This standardized comparison of major Alzheimer’s disease data modalities demonstrated that cognitive assessment provided the strongest single-modality prediction of 24-month progression from mild cognitive impairment to Alzheimer’s disease, while structural magnetic resonance imaging was the highest-performing biological modality. A multimodal model integrating all evaluated modalities achieved significantly higher discrimination than any single modality, including cognitive assessment, though this improvement was modest and required complete data from only one-fifth of the cohort. By evaluating all modalities and their combination within a common cohort using an identical analytical framework, this study provides a quantitative benchmark for comparing the prognostic value of routinely used clinical and biomarker data. The findings highlight the substantial predictive information contained in routinely collected cognitive assessments while clarifying the more limited incremental value of full multimodal integration under real-world data availability constraints. Future studies should validate these findings in independent, real-world clinical populations and evaluate calibration and clinical utility alongside discrimination.

## Data Availability

The data used in this study were obtained from the Alzheimer's Disease Neuroimaging Initiative (ADNI) database. ADNI data are publicly available to qualified investigators upon registration and approval through the ADNI data repository (https://adni.loni.usc.edu). The analysis code used to generate the results presented in this study will be made publicly available upon publication.

## List of Abbreviations

Aβ42: Amyloid-beta 42
AD: Alzheimer’s disease
ADNI: Alzheimer’s Disease Neuroimaging Initiative
APOE: Apolipoprotein E
AUC: Area under the receiver operating characteristic curve
CI: Confidence interval
CSF: Cerebrospinal fluid
FDG: Fluorodeoxyglucose
MCI: Mild cognitive impairment
MMSE: Mini-Mental State Examination
MRI: Magnetic resonance imaging
PET: Positron emission tomography
RAVLT: Rey Auditory Verbal Learning Test
ROC: Receiver operating characteristic

## Declarations

### Ethics approval and consent to participate

This study involved a secondary analysis of de-identified data obtained from the Alzheimer’s Disease Neuroimaging Initiative (ADNI) database. All ADNI participants provided written informed consent at their respective participating institutions, and the ADNI study received approval from the Institutional Review Board (IRB) at each participating site in accordance with the Declaration of Helsinki and applicable regulatory requirements. The present secondary analysis used publicly available de-identified data and did not involve direct contact with human participants.

### Consent for publication

Not applicable.

### Availability of data and materials

The data analyzed during the current study are available from the Alzheimer’s Disease Neuroimaging Initiative (ADNI) repository to qualified investigators upon application and approval. Information on data access is available at https://adni.loni.usc.edu. The code used to perform the analyses described in this study is available from the corresponding author upon reasonable request.

## Acknowledgements

Data collection and sharing for this project were supported by the Alzheimer’s Disease Neuroimaging Initiative (ADNI) (National Institutes of Health Grant U01 AG024904) and the Department of Defense ADNI (Award Number W81XWH-12-2-0012). ADNI is funded by the National Institute on Aging, the National Institute of Biomedical Imaging and Bioengineering, and through contributions from numerous public and private organizations. The grantee organization is the Northern California Institute for Research and Education, and the study is coordinated by the Alzheimer’s Therapeutic Research Institute at the University of Southern California. ADNI data are disseminated by the Laboratory for Neuro Imaging at the University of Southern California.

